# Automatic COVID-19 Detection from chest radiographic images using Convolutional Neural Network

**DOI:** 10.1101/2020.11.08.20228080

**Authors:** Sohaib Asif, Kamran Amjad

**Affiliations:** School of Computer Science and Engineering, Central South University Changsha, CHINA; School of Electronics and Information Engineering, Xi’an Jiaotong University Xian, CHINA

**Keywords:** COVID-19, Convolutional Neural Network, Biomedical imaging, Chest X-ray Radiographs

## Abstract

The global pandemic of the novel coronavirus that started in Wuhan, China has affected more than 50 million people worldwide and caused more than 1263,787 tragic deaths. To date, the COVID-19 virus is still spreading and affecting thousands of people. The main problem with testing for COVID-19 is that there are very few test kits available for a large number of affected or suspicious individuals. This leads to the need for automatic detection systems that use artificial intelligence. Deep learning is one of the most powerful AI tools available, so we recommend creating a convolutional neural network to detect COVID-19 positive patients from chest radiographs. According to previous studies, lung X-rays of COVID-19-positive patients show obvious characteristics, so this is a reliable method for testing patients, because X-ray examination of suspicious patients is easier than rt-PCR. Our model has been trained with 820 chest radiographic images (excluding data augmentation) collected from 3 databases, with a classification accuracy of 99.45% (training accuracy of 99.70%), sensitivity of 99.30% and specificity of 99.40 %, proved that our model has become a reliable COVID-19 detector.

## 1. Introduction

The novel coronavirus pandemic which originated in Wuhan, China in December 2019 has caused worldwide panic. The virus, which caused this COVID-19 pandemic, was called severe acute respiratory syndrome corona virus 2 or SARSCoV-2. It has been declared as a pandemic by the World Health Organization (WHO) on 11th March 2020 [1]. It infected more than 50 million people and died Charges are in thousands. New cases appear every day, this Brings huge challenges to healthcare professionals Find a cure and stop the spread. Even health care systems in developed countries It is about to collapse due to the increase in number. So far, there is no specific immunization or treatment for COVID-19. However, there are many continuous clinical preliminary evaluations of potential drugs. As of November 2, 2020, approximately 48,030,327 cases of infection have been confirmed in more than 216 countries, of which 1,223,171 have died, 34,465,814 have recovered, and 12,252,984 have mild and 88,358 critical cases were found [2, 3]. The distribution of COVID-19 cases between 28th August 2020 and 10th October 2020 are shown in the form of a bar graph in Fig 1. Their health requires patients in the intensive care unit. a lot of the country has activated a complete lockdown to prevent Disease spread. The current standard diagnosis of COVID-19 is the use of Real Time Polymerase Chain Reaction (rt-PCR). It is based on PCR technology [4], which is used to amplify and simultaneously detect or quantify a targeted DNA molecule on real-time basis. Reverse transcriptase real-time PCR formats can detect and quantify RNA molecules of the test organism in the sample on real time basis.

**Fig. 1:**
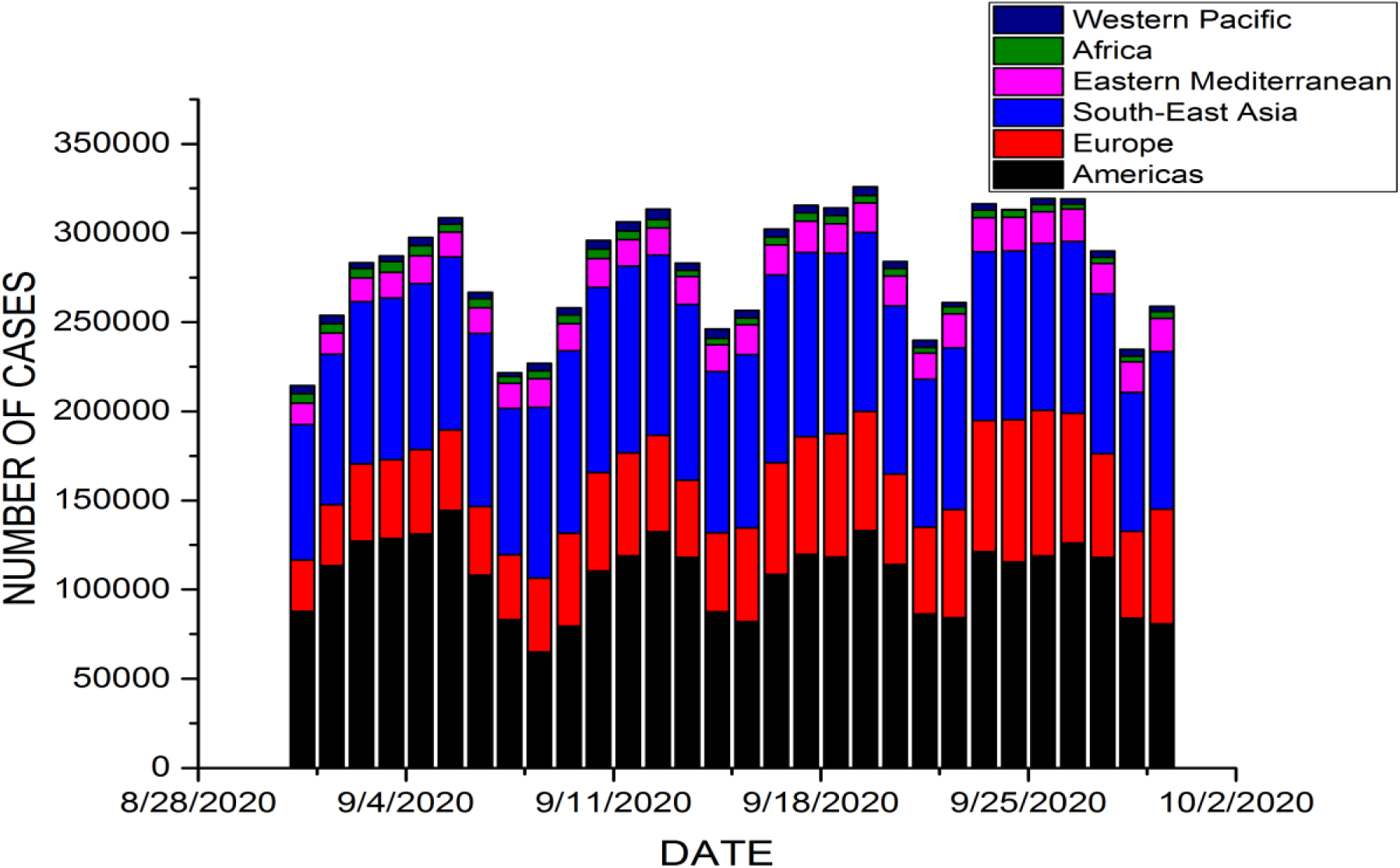
Bar graph of COVID-19 cases between 28th August and 10th October 2020.

It uses a different thermo cycler than the conventional PCR. But, the major problem for rt-PCR is that it is very expensive, 5-10 times more than the cost of conventional PCR. So, some hyper-endemic regions or countries are unable to provide the large number of rt-PCRs required for testing tens of thousands of suspected COVID-19 patients. This poses a possibility of some people to spread the virus without knowing they are infected. Radiographic examinations that are available in time are another major symptom tool of COVID-19. Most COVID-19 cases have comparable bright spots on radiographs, including intersecting, multifocal, ground-glass opacity with edges or spreading backwards, mainly in the lower projections in the early stages, and firm lungs in the early stages. [5-9] To address this problem of scarcity of apparatus, Computed Tomography or CT scan imaging has been used to detect the virus. Tao Ai et al. [10] reported a highest sensitivity of 97% based on detection from chest CT images of 1014 patients in Wuhan City, China. Fang et al. [11] reported the highest sensitivity of 98% based on chest CT images of 51 patients. Gozes et al. [12] employed deep learning methods for detection of COVID-19 positive patients using chest CT scan images. Shi et al. [13] developed and employed a machine learning based algorithm on a large COVID-19 CT scan images dataset for screening purpose. Wang et al. [14] developed a Residual CNN based neural network and achieved an accuracy of 92.4. Apostolopoulos et al. [15] employed transfer learning technique for classification of COVID-19, pneumonia and normal patients’ chest X-rays and the best accuracy achieved was 96.78%. [16] proposed a framework based on Capsule Network, known as the COVID-CAPS, for COVID-19 identification using X-ray images. The proposed COVID-CAPS achieved 95.7% accuracy, 90% sensitivity, 95.8% specificity, and 0.97Area Under the Curve (AUC). Ayrton [17] used a small dataset of 339 images for training and testing using ResNet50 based deep transfer learning technique and reported the validation accuracy of 96.2%

However, there are drawbacks of CT scan imaging which includes the considerably longer time required for the test than X-ray imaging [18] Also, availability of high-quality CT scanners may be scarce in certain underdeveloped regions, which can make the timely detection of COVID-19 patients very difficult or impossible. On the flip side X-ray diagnosis is the most common imaging technique and it is widely used for clinical care and epidemiological studies. So, using X-ray imaging for COVID-19 detection can speed up the timely screening of patients. Also adding to the fact that since March 2020, the amount of publicly available X-ray images of COVID-19 positive patients has increased, it enables us to study the medical images for possible patterns for the automatic diagnosis of corona virus infection.

Deep Learning applications have evolved over the past few years and the problem at hand seems to be something that can be addressed using the deep learning techniques for highly accurate screening of COVID-19 patients. Deep Learning is a machine learning technique that teaches a computer how to learn by examples, something which humans often do. So artificial “neural networks” are built so that the computer can learn through the examples (dataset) we provide. Deep Learning is commonly used for object detection, classification, segmentation, lesion detection problems in medical imaging. These datasets can be obtained from biomedical imaging techniques like Magnetic Resonance Imaging (MRI), radiographic image (X-Rays), Computed Tomography (CT), ultrasonography, endoscopy among others. Also, images of slides like histopathological and cytological slides are used as data for cancer detection based deep learning models. These models help in near perfect detection of diseases like breast cancer, diabetes, tumor, skin infections, etc. A Convolutional Neural Network (CNN) is a special application of deep learning that is used mostly for recognition, classification and segmentation purposes. As the name suggests, there must be at least one layer that performs convolution operation instead of general matrix operations. This makes the neural network different from general neural networks. CNN’s are actually perceptron’s with multiple layers and simplified architecture. They have layers like activation, convolution, softmax, identity, classification etc. The use of artificial Intelligence will aid radiologists in detecting COVID-19 positive patients which is highly desired since it might be difficult for them to visually analyze the X-rays and distinguish patients.

So, motivated by the dire need to develop a way to help fight the disease by detecting patients who are COVID-19 positive and to help the research community who provided with the open source databases, we developed COV-19Net, a Convolutional Neural network tailored to distinguish between COVID-19 positive and normal patients through chest X-Ray images. This can aid the medical practitioners fighting against the COVID-19 pandemic by detecting cases faster. Examples of chest radiographic images of a COVID-19 positive patient and a tested negative patient are shown in Fig 2a and Fig 2b respectively.

**Fig. 2:**
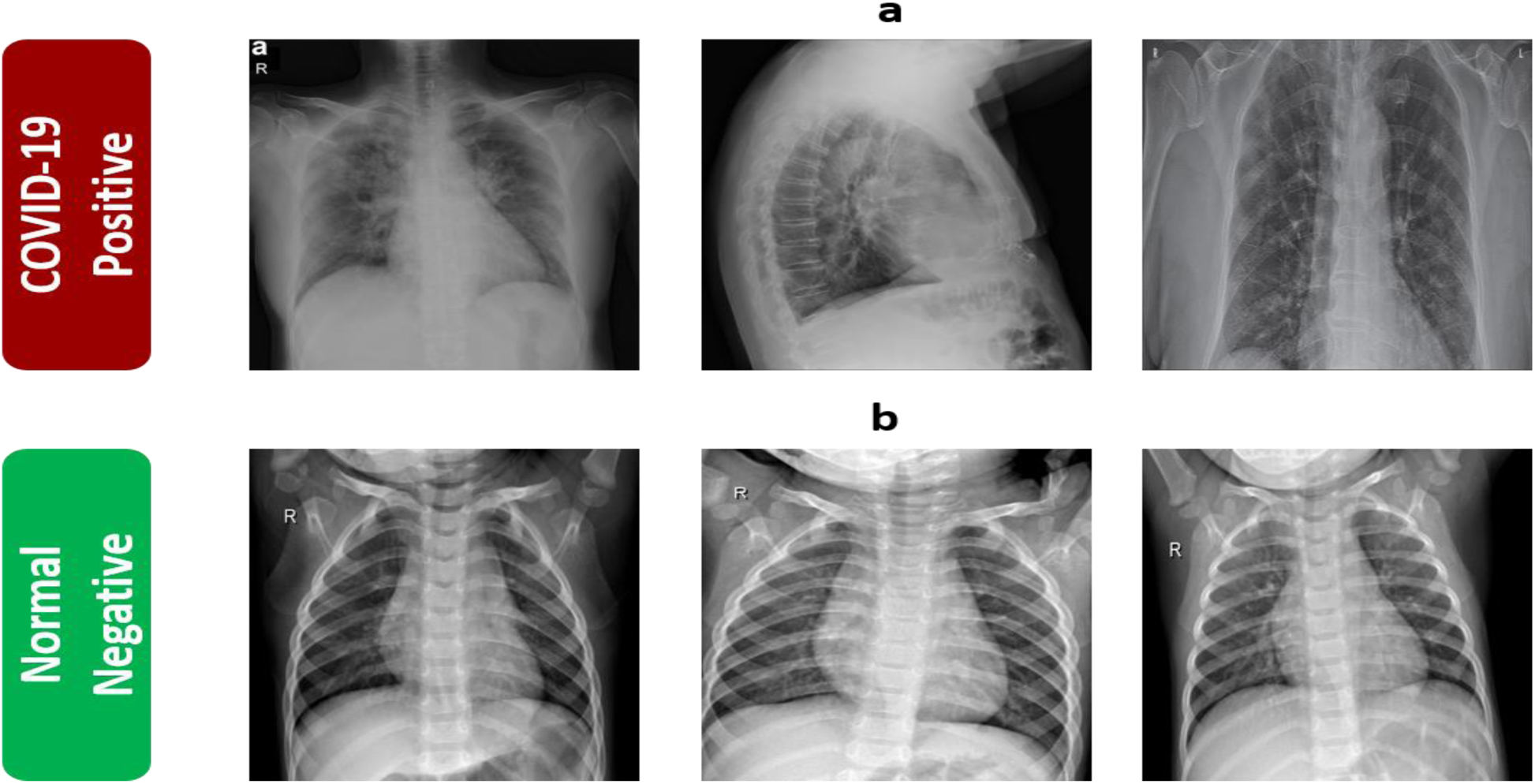
(a) Chest radiographic image of a COVID-19 positive patient (b) Chest radiographic image of a COVID-19 negative patient.

**Fig. 3:**
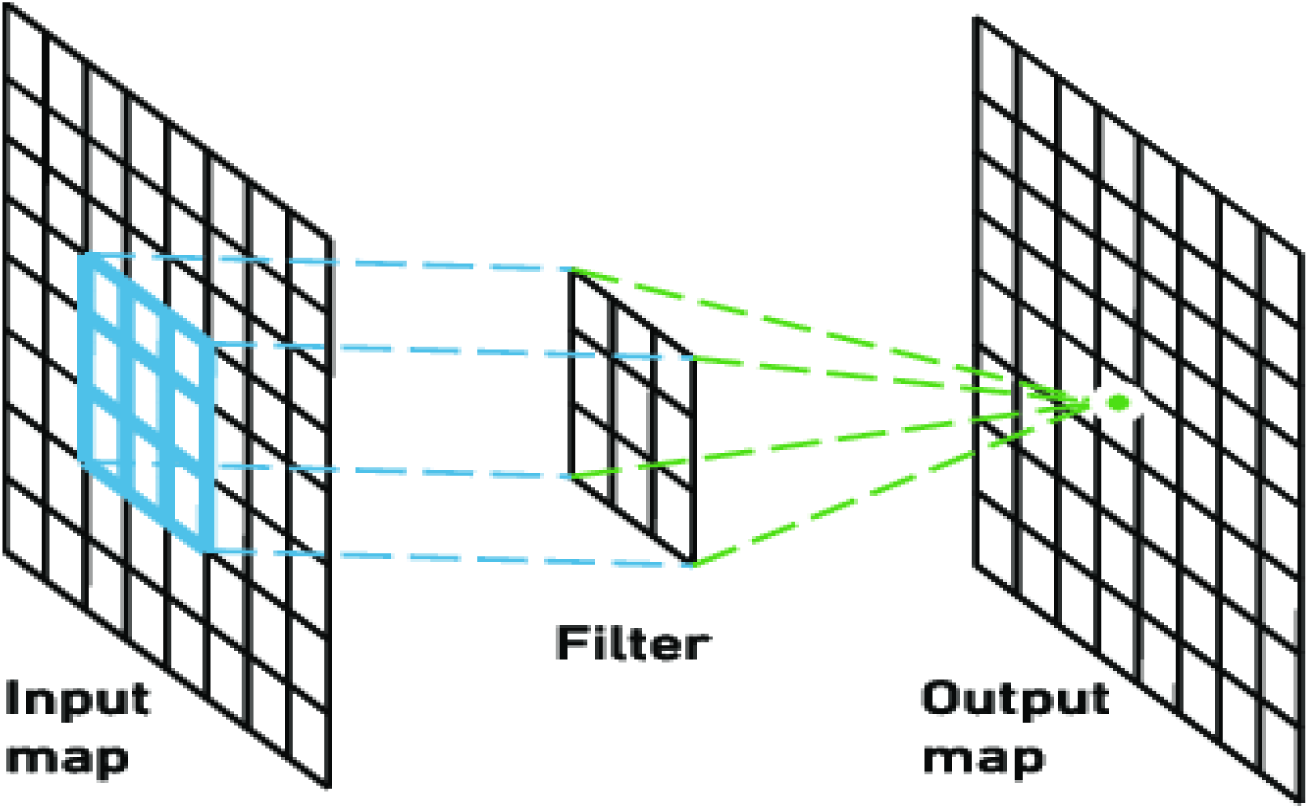
Example of Convolutional Layer

The rest of the paper has been organized into sections. Section 2, Materials and Methods, describes the architecture of the neural network used, the dataset and the implementation procedures. Section 3, Results and Discussions describes in detail the results obtained from the experiments and finally Section 4, Conclusions and Future Works, draws the conclusions from the research and possibility of future work on the proposal.

## 2. Convolutional Neural Network (CNN)

Convolutional neural network (CNN) [19] is a network architecture for deep learning. CNN is deep Artificial neural network is mainly used for image classification, clustering them through similarity and performing Object recognition in the scene. CNN consists of one or more convolutional layers, and then follow Like one or more fully connected layers in a standard multilayer neural network, it learns directly from the image. A CNN can be trained to perform image analysis tasks, including classification, target detection, segmentation and images processing.

This whole process of creating CNN is done by three layers

- Convolutional Layer
- Pooling Layer
- Fully Connected Layer.

### 2.1 Convolutional Layer

Convolutional layers are a key component of convolutional neural networks, and are always at least their first layer. Its purpose is to detect whether there is a set of features in the image received as input. This is done through convolution filtering: the principle is to “drag” the window representing the feature on the image, and calculate the convolution between the feature and each part of the scanned image. Then think of features as filters: in this context, the two terms are equivalent. The convolutional layer therefore receives several images as input and uses each filter to calculate the convolution of each image. The filter corresponds exactly to the function we want to find in the image. We obtain a feature map for each pair (image, filter), which tells us the location of the feature in the image: the higher the value, the more similar the corresponding location in the image is to the feature.

### 2.2 Pooling Layer

Usually we will add a pooling layer among two convolutional layers in a standard convolutional neural network structure. The usage of pooling layer is to rapidly decrease the size of data in the middle layers so that we can reduce the amounts of calculations which results from a smaller number of parameters and weights of each neuron. Through this approach, the problem of overfitting can also be efficiently controlled. The pooling layers can work independently with the MAX function, which can form the well-known max-pooling layers. One of the most popular instances of a pooling layer is layer with filters of size 2 * 2. This kind of pooling layer with stride 2. The depth, height as well as width of input will be reduced to the half of their original size, and thus the total size of output can be reduced to just 25% of the original size.

### 2.3 Fully Connected Layer

This type of layer receives an input vector and produces a new output vector. To do this, it applies a linear combination and then possibly an activation function to the input values received. It takes the output of the previous layers, “flattens” them and turns them into a single vector that can be an input for the next stage.

## 3. Material and Methods

In this section we will describe the datasets used, the architecture of the COV-19Net and the Implementation Procedure in the creation of the COV-19Net.

### 3.1 Collection of Dataset

For the training and evaluation of the COV-19Net we have collected chest radiographic images from 3 open source databases: 1) GitHub open source dataset by Chowdhury et al. [20] 2) Dataset from GitHub by Cohen et al. [21] 3) Positive radiographic images (CXR and CT) were carefully chosen from Italian Society of Medical and Interventional Radiology (SIRM) [22] COVID-19 DATABASE. All the images obtained were resized to 64×64×3 pixel size. We used 410 images of COVID-19 positive patients and 410 images of COVID-19 negative patients. Data augmentation was employed on the images to increase the data and the techniques used were rescaling, vertical flipping, horizontal flipping, rotating, zooming, intensity shift and shear intensity shift (shear angle in counter-clockwise direction) which gave us a dataset of total 2870 images of each class (COVID positive and negative), i.e. a total of 5740 images. The training images were 70% of total images and the rest 30% comprised the validation set. There is only a very small amount of data available through more than a couple of million people have been tested and diagnosed. This suggests the need for more open source data for building even better networks for the detection of COVID-19 through Artificial Intelligence and for studying trends in the X-Ray images.

### 3.2 COV-19Net Architecture

A CNN network is composed of 3 types of layers namely, Convolution, Pooling and Dense Layer or Fully Connected Layers. The convolution layer has filters of a specified size with learnable parameters and these filters are convolved or mathematically speaking, cross-correlated with the input image (which is a matrix too). The output is another matrix. Now the shape of the output matrix depends on the input matrix size, the padding and the number of filters used for convolution. Padding means adding a row and column to the input matrix all of whose components are zeroes. This is done to make the height and width of the output matrix the same as that of the input matrix. Padding, hence is of two types ‘same’ and ‘valid’. ‘Valid’ padding essentially means no padding at all, while padding type ‘same’ means the padding is done such that the height and width of the input and output matrices are the same. The depth or number of channels of the output matrix is the same as the number of filters used for the convolution. Striding means the number of pixels to be shifted by the kernel for operation (convolution or pooling). For example, if stride is 1, the kernel is shifted one pixel at a time for convolution or pooling operation. Pooling Layers are usually of two types namely, average pooling and max pooling. It is basically a dimensionality reduction or feature extraction step. In Max Pooling the kernel size is swept through the input matrix and the max of the numbers inside the matrix is selected and put in the output matrix, or in Average Pooling the average of the numbers is calculated inside the kernel size and put in the output matrix. In both cases normally the striding is done according to the size of the pool kernel. An activation function is used just before the Pooling operation, like sigmoid activation, tanh activation or ReLU activation or softmax activation for multiclass problem. A simple example of max and average pooling is shown in Fig 4.

**Fig. 4:**
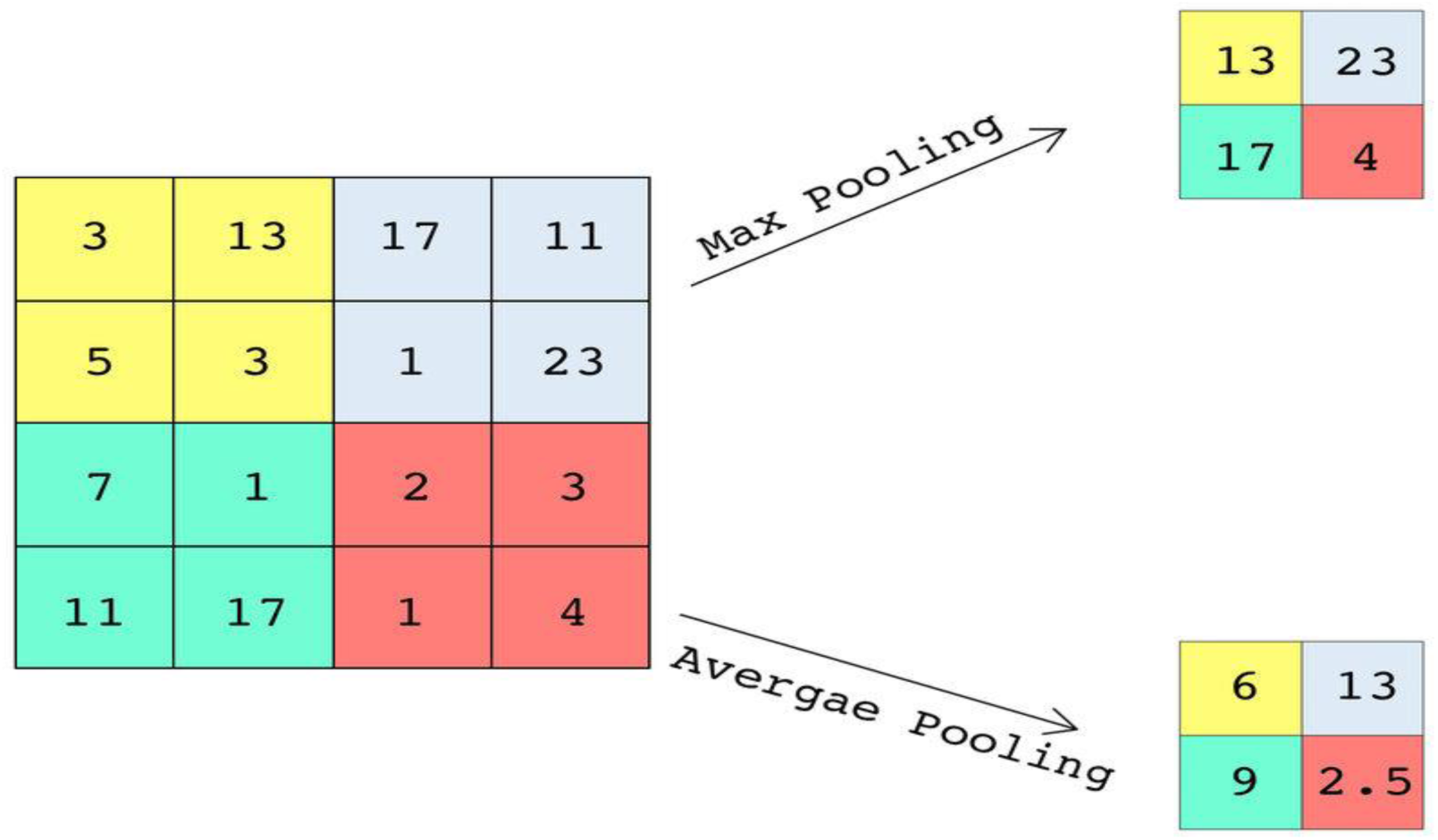
Example of Max and Average Pooling.

After the final Convolution or Pooling operation, the matrix, say, is of shape (l, m, n), it is flattened out that is it is converted to a vector of shape (l x m x n, 1). Next these are connected to the final set of layers of the CNN called the Dense Layers or the Fully Connected layers because all neurons of the flattened layer are connected to all neurons of the Fully Connected Layers. These layers are essentially the same as in other types of neural networks. The final fully connected layer is also the final layer of the CNN and has number of neurons as number of outputs required. For multi-class problem, the number of outputs is equal to the number of classes and the activation is softmax. For binary classification problem, the number of outputs is 1 in case of sigmoid activation and 2 in the case of softmax activation.

The COV-19Net Convolutional Neural Network is composed of 3 convolution layers, 2 max pooling layers, and 2 fully connected Dense Layers and is as shown in the flowchart of Fig 5. The layer by layer details of COV-19Net are presented as follows:

➢ Layer 1: This is a convolution layer where the input image of size 64×64×3, is convolved with 32 filters of kernel size 3×3, without any padding, stride=1 and ReLU activation.
➢ Layer 2: This is a 2D Max Pooling layer of shape 2×2, without padding and strides = 2.
➢ Layer 3: This is the second convolution layer with 32 filters of kernel size 3×3, without any padding, stride=1 and ReLU activation.
➢ Layer 4: This is a 2D Max Pooling layer of shape 2×2, without padding and strides = 2.
➢ Layer 5: This is the third and final convolution layer of 32 filters and kernel size of 3×3, without any padding, stride=1 and ReLU activation. The output of this layer is flattened out to form a 4608×1 vector
➢ Layer 6: This is the first fully connected layer or dense
➢ Layer of 128 units and ReLU activation.
➢ Layer 7: This is the second fully connected layer and final layer of the COV-19Net consisting of 1 unit with sigmoid activation function.

**Fig. 5:**
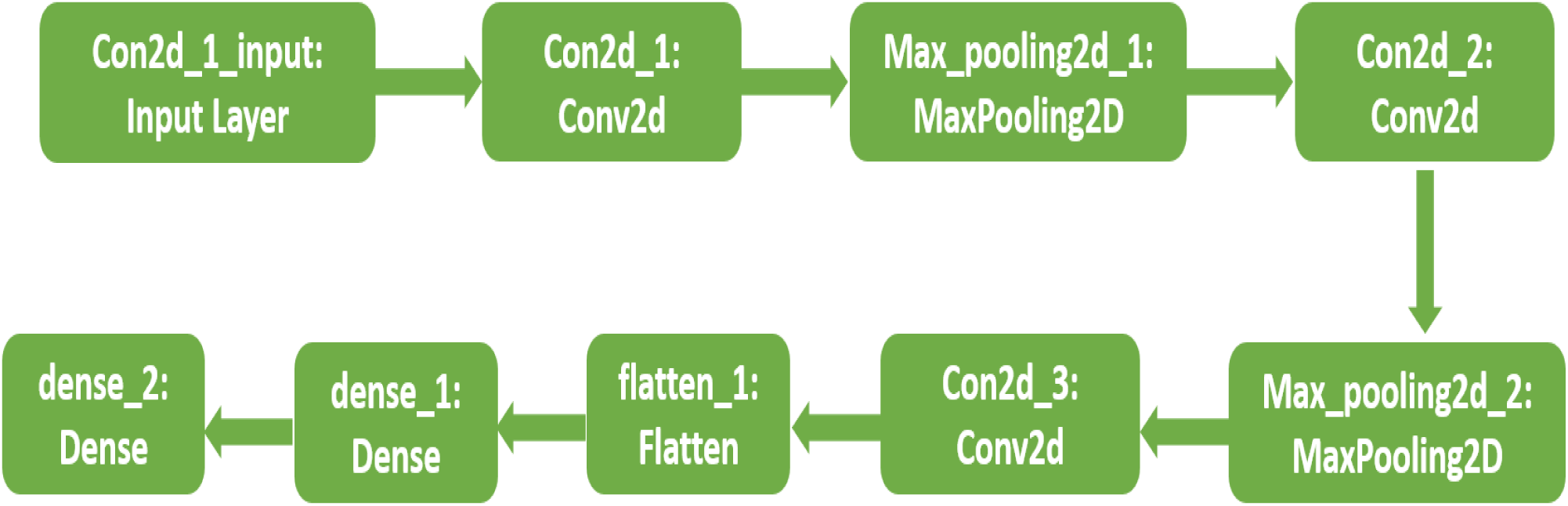
Flow chart diagram of COV-19Net Architecture.

The architecture diagram of COV-19Net is shown is Fig 6.

- Model Type: Sequential
- Total parameters: 609,473
- Trainable parameters: 609,473
- Non-trainable parameters: 0

**Fig. 6:**
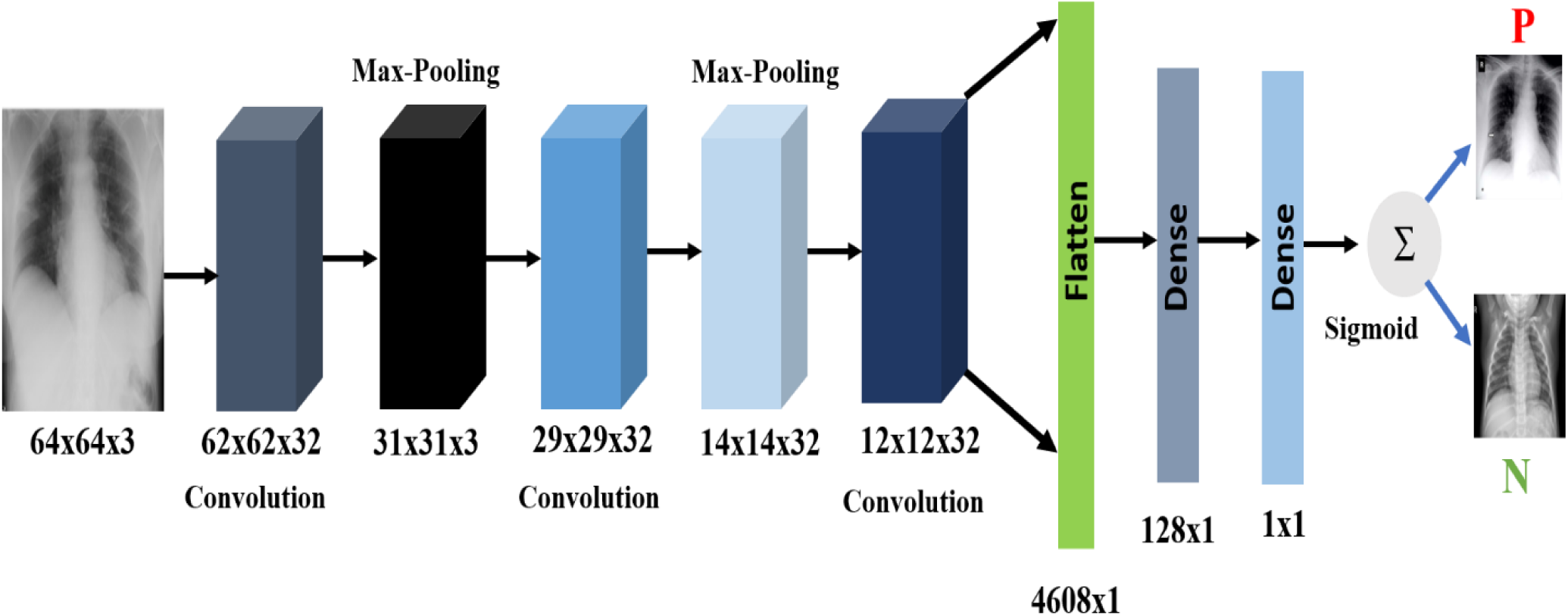
Architecture diagram of COV-19Net.

### 3.3 Implementation

The network was trained using our dataset by giving random weight initialization. The inputs were of shape 64×64×3. We used Adam optimization [23] and the hyper-parameters used were constant learning rate of 0.001, batch size of 32, steps per epoch 100 and number of epochs 20. Further we implemented data augmentation due to shortage of resources to train our model and the types of augmentation used were: rescaling, rotating, zooming, vertical flip, horizontal flip, intensity shift and shear intensity shift (shear angle in counter-clockwise direction). The proposed COV-19Net model was implemented through the Keras framework using TensorFlow[24] backend.

## 4. Results and Implementation

The parameters measured for the proposed COV-19Net are accuracy, specificity, precision, recall or sensitivity and f1 score which are calculated for both the training set and the validation set. First, we need to define the terms True Positive (TP), False Positive (FP), True Negative (TN) and False Negative (FN) for defining the aforementioned parameters. In a binary classification model let there be two classes positive and negative. If the model classifies an image to belong to the positive class and the image actually belongs to the positive class, meaning the classification is correct, it is called True Positive. If the model classifies an image to belong to the negative class whereas actually the image is part of the positive class, it is called False Positive. If the model classifies an image to belong to the negative class and the image actually belongs in the negative class it is classed True Negative. If the model classifies an image to belong to the positive class but actually the image is part of the negative class, it is called to be False Negative.

Now the formulae for Accuracy, Specificity, Precision, Recall or Sensitivity and F1 Score are given as follows:

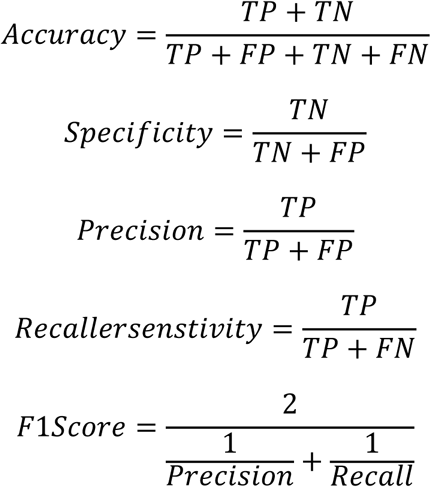

The model was trained for 200 steps epoch for 20 epochs. The values of all the mentioned parameters and the loss obtained after Epoch 20 on training set and validation set are provided in Table 2. The values of accuracies obtained on the Training set and the Validation set being so close to each other suggests low variance and the high value of accuracy suggests low bias. So, our COV-19Net gives almost impeccable results and can be thought of as an ideal model for use by the medical practitioners for faster detection of COVID-19 positive patients from chest radiographic image. The plots for model losses and model accuracy based on number of epochs are shown in Figure 7 and 8 respectively.

**TABLE I:** Output shape and number of parameters for each layer.

| Layer Type | Number of filters/units | Kernel Size | Strides | Padding Type | Output Shape | Number of Parameters |
| --- | --- | --- | --- | --- | --- | --- |
| **Convolution** | 32 filters | 3x3 | 1 | Valid | 62x62x32 | 896 |
| **Max Pool** | 32 filters | 2x2 | 2 | Valid | 31x31x32 | 0 |
| **Convolution** | 32 filters | 3x3 | 1 | Valid | 29x29x32 | 9248 |
| **Max Pool** | 32 filters | 2x2 | 2 | Valid | 14x14x32 | 0 |
| **Convolution** | 32 filters | 3x3 | 1 | Valid | 12x12x32 | 9248 |
| **Flatten** | 4608 units | - | - | - | 4608x1 | 0 |

**TABLE II:** Results after training the COV-19Net

| Set | Accuracy | Specificity | Precision | Recall/Sensitivity | F1 Score | Loss after final Epoch |
| --- | --- | --- | --- | --- | --- | --- |
| **Training** | 99.70% | 99.68% | 99.89% | 99.91% | 99.70% | 0.0035 |
| **Validation** | 99.45% | 99.40% | 100% | 99.30% | 99.43% | 0.0018 |

**Fig. 7:**
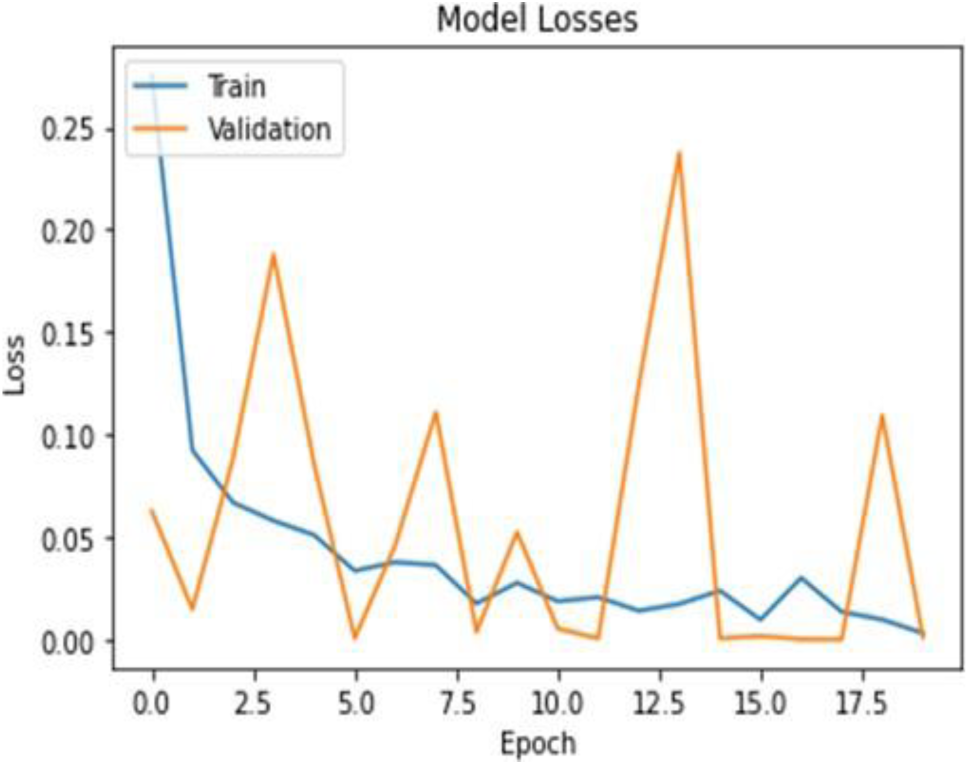
Training and validation losses and accuracy plot

**Fig. 8:**
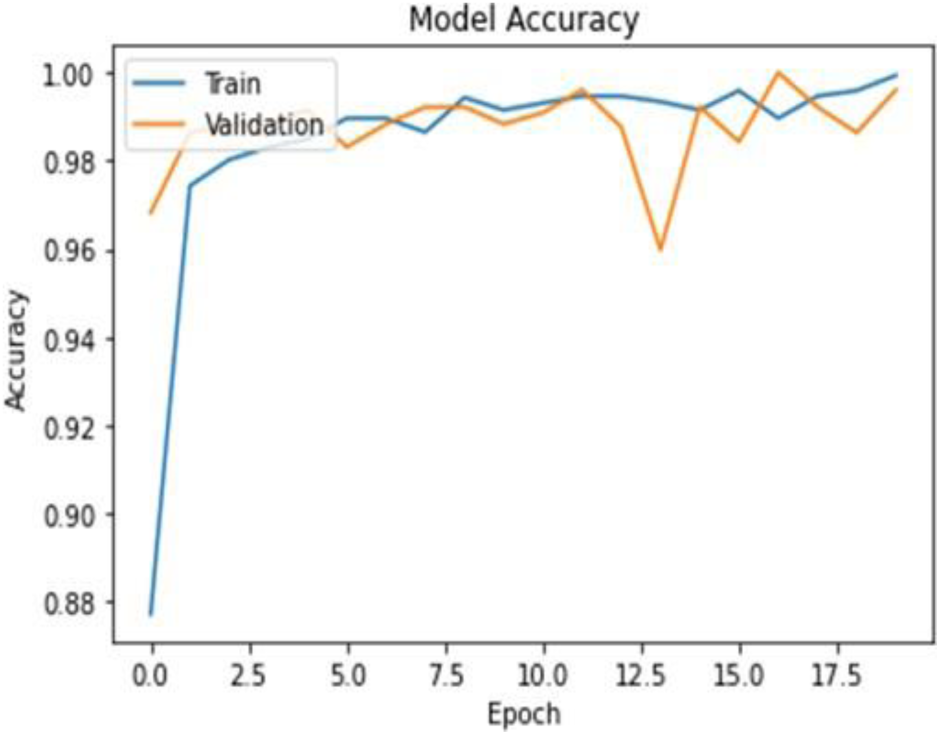
Training and validation losses and accuracy plot

Zhang et al. [25] achieved best sensitivity and specificity of 90% and 87.84% on their neural network, Narin et al. [26] used transfer learning on a variety of models and achieved best mean accuracy of 98% on ResNet50 model, and Apostolopoulos et al. [15] also employed transfer learning and achieved best accuracy of 96.78% on VGG-19 network. To summarize, a comparison of other existing models and the proposed COV-19Net is shown in Table 3. From Table 3, it is clear that the proposed COV-19Net outperforms all existing models and is a very reliable model for COVID-19 detection. Along with the higher accuracy, specificity, sensitivity and F1 scores than all existing models, the other advantages of the COV-19Net include that the model is an end-to-end model that is there is no feature extraction or feature selection steps; the model uses more widely available lung X-Ray images for predictions which is a more efficient method that RT-PCR tests since enough test kits are not available for the rapidly growing population of COVID-19 infected patients.

**TABLE III:** Comparison of the results obtained from the proposed network as compared to the existing networks.

| Methods | Best Accuracy |
| --- | --- |
| Narin et al. [26] | 98.5% |
| Bukhari et al. [27] | 98.18% |
| Apostolopoulos et al. [15] | 96.78% |
| Wang et al. [14] | 92.4% |
| **Our Proposed COV-19Net Model** | **99.45%** |

## 5. Conclusion and Future Work

In this research, we proposed the convolutional neural network COV-19Net as an effective computer-aided diagnosis method for COVID-19 detection. Since this method uses chest X-ray images for prediction and is an end-to-end system, this method is also more effective, which means there are no feature extraction and/or selection steps. It is vital to quickly detect infected patients to take necessary actions and preventive measures, and in the rapidly growing population of infected patients, fighting the pandemic becomes particularly important. If more clinical data are available, the COV-19Net model can be further verified and its effectiveness can be better judged. We hope that our model has high accuracy and sensitivity, as well as cost-effective and fast-implemented procedures, so it can help society fight this pandemic and reduce suffering. In addition, the model can also help clinicians detect the difference in X-rays between COVID-19 positive and COVID-19 negative patients and help them fight the virus. Our future work will focus on collecting X-ray data from more patients and incorporating it into our model for better performance.

## Data Availability

GitHub open-source dataset by Chowdhury et al.
The dataset from GitHub by Cohen et al.

